# Tocilizumab Reduces Hypercoagulation in COVID-19 – Perspectives from The Coagulation and Immunomodulation Covid Assessment (Coag- ImmCovA) Clinical Trial

**DOI:** 10.1101/2024.05.31.24308108

**Authors:** Lou M. Almskog, Anna Sjöström, Jonas Sundén-Cullberg, Apostolos Taxiarchis, Anna Ågren, Sara Freyland, Madeleine Börjesson, Agneta Wikman, Carl Magnus Wahlgren, Michael Wanecek, Jan van der Linden, Jovan Antovic, Jon Lampa, Maria Magnusson

## Abstract

**Background:** Despite medical interventions, COVID-19 continues to persist at pandemic proportions. A hypercoagulation state was rapidly observed in the severely ill, and the incidence of thromboembolic events remains elevated. Interleukin inhibitors have demonstrated positive effects on the hyperactivation of the immune system in COVID-19, with the interleukin-6 inhibitor tocilizumab showing promising results in reducing mortality. Nevertheless, the impact of interleukin inhibitors on the coagulation system remains incompletely understood.

**Methods:** In this clinical trial conducted in Stockholm, Sweden, interleukin inhibitors, namely anakinra (ANA) or tocilizumab (TOCI), were randomly administered in addition to standard care (SC) to hospitalized patients with COVID-19. A control group received only SC. The primary outcome sought to measure effects on global hemostasis, as indicated by changes in functional coagulation tests, specifically Rotational Thromboelastometry (ROTEM) or Overall Hemostatic Potential (OHP), visualized through scanning electron microscopy images. Secondary outcomes included effects on conventional coagulation laboratory tests.

**Results:** The study enrolled 74 patients who were randomized to receive either ANA or TOCI in addition to SC, or SC alone. In the TOCI group, ROTEM variables exhibited less hypercoagulation after 29 days compared with ANA or SC treatment groups, characterized by prolonged clot formation time and decreased clot firmness. OHP decreased, but there were no significant differences among the three treatment groups. Plasma fibrinogen levels, initially elevated, decreased significantly in TOCI recipients over time.

**Conclusion:** Tocilizumab treatment demonstrated a significant reduction of hypercoagulation in hospitalized COVID-19 patients, by improvements in both global coagulation tests and conventional laboratory tests, in comparison with anakinra or SC alone. This finding underscores the significance of tocilizumab as a viable treatment option in severe COVID-19 cases, with the potential to decrease thrombosis incidence.

## Introduction

Coronavirus Disease 2019 (COVID-19) remains an ongoing pandemic, with over 770 million confirmed cases, and 7 million deaths reported ^1^. In individuals with severe COVID-19, a dysregulated hyperactivation of the immune system is frequently observed, accompanied by elevated serum levels of cytokines and acute phase reactants ^2,3^. These cytokines play a crucial role in activating the extrinsic coagulation cascade and inhibiting fibrinolysis, resulting in microvascular thrombosis, posing potential risks of organ dysfunction ^4^.

Pro-inflammatory cytokines, such as interleukin 1 (IL-1) and interleukin 6 (IL-6), are pivotal mediators of acute inflammation, inducing the expression of various human cell proteins ^5,6^. Prior to the pandemic, IL-1 receptor inhibitor anakinra and IL-6 receptor inhibitor tocilizumab demonstrated efficacy in reducing hyperinflammation in autoinflammatory and systemic inflammatory conditions ^7,8^. The early focus on cytokines as treatment targets in severe COVID-19 led to clinical trials that demonstrated decreased mortality with cytokine blockade in severely ill patients ^9,10^.

COVID-19 is characterized by a hypercoagulable condition, with disturbances in the hemostatic balance contributing to a highly prothrombotic state ^11,12^. High thrombosis incidence has been observed in both the arterial and venous circulation of COVID-19 patients, accompanied by elevated D-dimer and plasma fibrinogen levels ^13–17^.

Recent studies on global coagulation tests in COVID-19 have further detailed clot strength and fibrinolysis abnormalities. Whole blood Rotational Thromboelastometry (ROTEM) revealed markedly procoagulative profiles, reflected by faster clot activation rate (clot formation time, CFT) and increased clot firmness (maximum clot firmness, MCF) ^18,19^. Similarly, Lee et al. found elevated fibrin generation (Overall Coagulation Potential (OCP) and Overall Hemostatic Potential (OHP)) and impaired fibrinolysis (Overall Fibrinolytic Potential (OFP)), associated with poor prognosis and increased morbidity ^20^. Confocal microscopy studies by Whyte et al. revealed a denser fibrin network with more branching in COVID-19 patients, and Boknäs et al. demonstrated impaired fibrinolysis in COVID-19 through analysis of fibrin clot structure ^21,22^.

The IL-1 family, pivotal in local and systemic responses to infection and inflammation, includes IL-1α, IL-1β, and the highly selective endogenous receptor antagonist IL- 1Ra. Anakinra (Kineret®, Swedish Orphan Biovitrum, Solna, Sweden), blocks the activity of both IL-1α and IL-1β with a short terminal half-life of 4 to 6 hours ^7,23,24^. IL- 6, a pleiotropic cytokine with crucial functions in inflammatory processes, plays a key role in connecting the immune and coagulation systems ^25^. Tocilizumab (RoActemra®, Roche, Basel, Schweiz), a humanized IL-6 receptor antibody completely inhibiting the binding of IL-6, has been extensively used in rheumatologic diseases and is characterized by a non-linear pharmacokinetic profile ^26^. In this context, drug elimination is influenced by factors that cause deviations from a simple linear relationship, and adjustments in dosage do not result in proportional changes in drug levels ^8^.

The dysregulated hyperactivation of the immune system in severe COVID-19, leading to high cytokine levels, may induce a state of hypercoagulation, resulting in clot formation and organ damage ^27^. In this state of “immunothrombosis” ^27,28^, anticoagulant treatment alone may not suffice, necessitating simultaneous reduction of the immunological impact. Treatment with anakinra and tocilizumab could potentially reduce coagulation activation and thrombosis risks in COVID-19 patients.

Our objective was to assess, in a clinical trial involving patients with severe COVID- 19, whether interleukin inhibitors anakinra or tocilizumab have significant effects on global hemostasis tests and conventional coagulation laboratory tests, indicative of reduced hypercoagulation patterns compared to standard care alone.

Our pre-registered hypotheses (AsPredicted Submission #129368, AsPredicted.org), were as follows:

**A)** In patients experiencing respiratory distress caused by COVID-19 and receiving interleukin antagonists anakinra or tocilizumab in addition to SC, global hemostasis tests and coagulation laboratory analyses will exhibit less hypercoagulation compared to patients receiving SC alone.
**B)** These effects will be more pronounced after tocilizumab treatment compared to anakinra treatment.

## Materials and methods

### Trial design

The Coag-ImmCoVA study is a supplementary investigation stemming from the Immunomodulation-CoV Assessment Multi-center (ImmCoVA) Study, which took place at four sites in Sweden from June 2020 to March 2021 ^29^. In the ImmCoVA Study, a total of 77 eligible participants were randomly assigned in a 1:1:1 ratio to one of three arms: interleukin-1 antagonist anakinra (ANA arm), interleukin-6 antagonist tocilizumab (TOCI arm), both in addition to standard care (SC), or standard care alone (see Fig. 1). The inclusion of patients involved block randomization with stratification based on study site, sex, and age. Further details regarding the randomization process, data collection methods, and patient follow-up procedures are available in the main analysis of the ImmCoVA Study.

**Figure 1.**
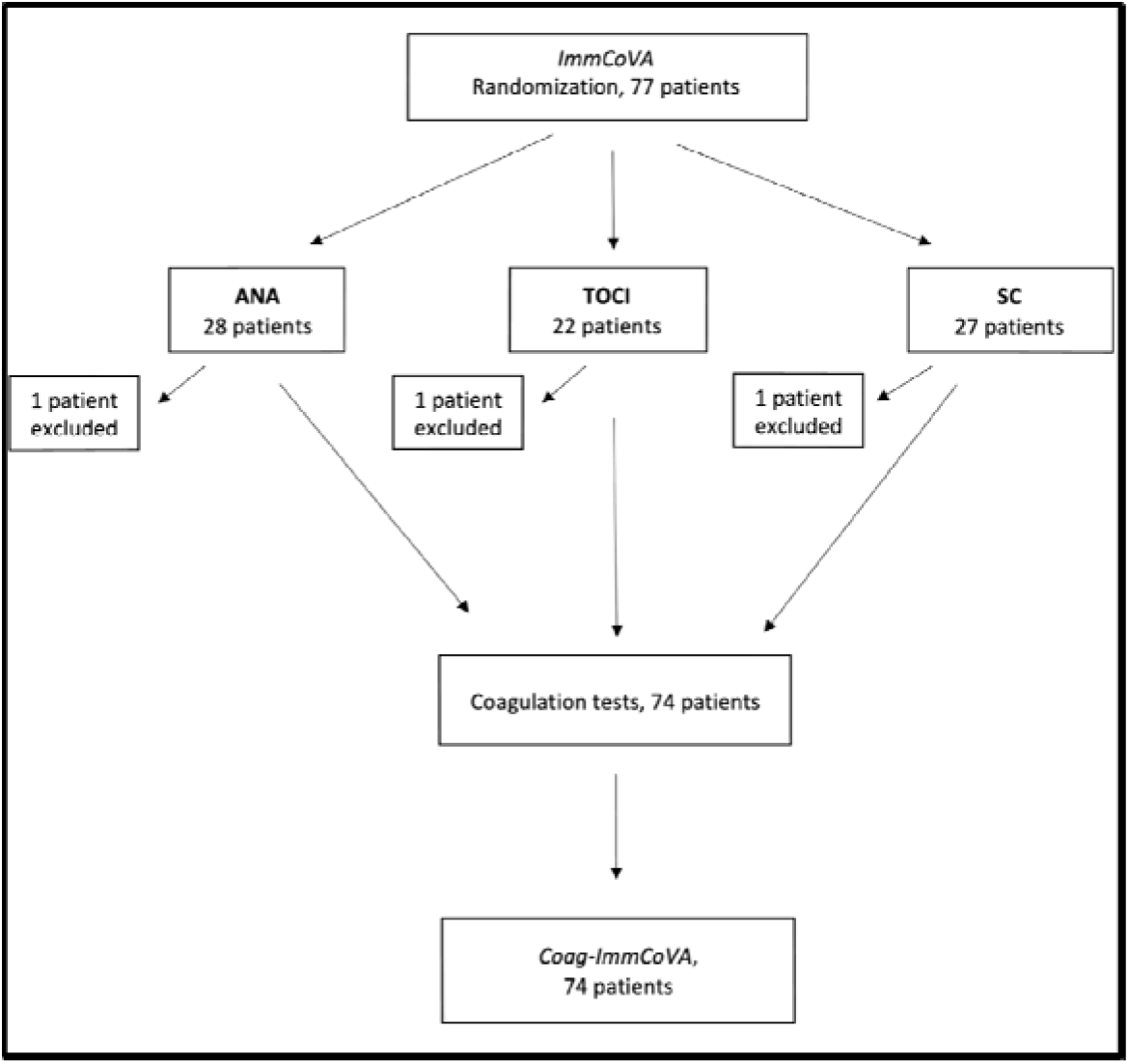
Study flowchart. Three patients were excluded after randomization due to hospitalization at other sites.

The Coag-ImmCoVA study comprised the Karolinska University cohort from the ImmCoVA study, encompassing 74 patients. Within this cohort, 27 patients were allocated to the ANA arm, 21 patients to the TOCI arm, and 26 patients to the SC arm, as illustrated in Figure 1. Ethical approval for the study was obtained from the Swedish Ethical Review Authority (D-nr 2020-01973, 2020-05709), and the original trial protocol is accessible on clinicaltrials.gov (ImmCoVA, Id: NCT04412291).

### Inclusion and exclusion criteria

Hospitalized adults (≥ 18 years of age) at Karolinska University Hospital were assessed for eligibility based on the following criteria: confirmation of SARS-CoV-2 infection through polymerase chain reaction (PCR) or public health assay test conducted within 3 days before enrollment; a duration of at least 7 days from the onset of symptoms to inclusion; a requirement for oxygen supply of ≥ 5 L/min for a minimum of 8 hours to maintain a SpO2 of ≥ 93% (or a shorter duration in cases of acute deterioration and a need of > 10 L/min to maintain SpO2 ≥ 93%); CRP > 70 mg/L; Ferritin > 500 µg/L; fulfillment of at least two out of three laboratory test criteria, including lymphocytes < 1 x 10^9^/L, D-dimer ≥0.5 mg/L FEU, and Lactate Dehydrogenase ≥ 8 µkat/L. Written informed consent was obtained from all participants.

Patients meeting any of the following criteria were excluded: mechanical ventilation during the current hospital stay; severe renal dysfunction (eGFR < 30 ml/min); confirmation of acute systemic non-SARS-CoV-2 infection through blood cultures; presence of chronic liver disease; uncontrolled hypertension (systolic BP > 180 mmHg or diastolic BP > 110 mmHg); history of autoimmune or inflammatory disease other than rheumatoid arthritis; history of stem-cell or solid organ transplantation; previous tuberculosis, HIV or acute/chronic viral hepatitis; history of gastrointestinal ulceration or diverticulitis; known hypersensitivity to the study drugs; presence of any of the following laboratory findings at inclusion: absolute neutrophil count < 2 x 10^9^/L, aspartate aminotransferase or alanine aminotransferase > 5 times the upper normal value, platelet count < 100 x 10^9^/L; pregnancy or breastfeeding; of reproductive age and not willing to use contraceptive methods until 3 months after last dose of the study drug; expected survival < 48 hours from inclusion; expected overall survival due to comorbidities < 3 months; or participation in any clinical research study evaluating an investigational product (excluding Remdesivir) within the last 3 months.

### Intervention/Treatments

Eligible patients underwent treatment allocation through randomization and were assigned in a 1:1:1 ratio to receive SC only, or, in addition to SC, either anakinra or tocilizumab. Supportive SC adhered to national guidelines and treatment protocols. Patients allocated to the ANA arm received anakinra, administrated at a dose of 100 mg intravenously every 6 hours for 7 days from baseline. Patients in the TOCI arm received tocilizumab as a single dose of 8 mg/kg intravenously, up to maximum 800 mg, at baseline. In cases of renal dysfunction (eGFR 30-59 ml/min), anakinra doses were reduced to 100 mg every 12 hours. Steroids were permissible if already taken before hospitalization at a daily dose < 10 mg prednisolone or equivalent, or as part of SC ≤ 5 days before inclusion. All drugs were allowed during the study period, except for disease-modifying antirheumatic drugs (DMARDs), with no administration in the past 30 days. Antibiotic prophylaxis was administrated in the intervention arms for 7 days (ceftriaxone, cefotaxime, or clindamycin and ciprofloxacin). Low molecular weight heparin (LMWH) was administered according to standard care protocols, with anti-FXa levels measured to assess anticoagulant effects.

### Clinical outcomes

The primary objective was to observe significant and measurable effects on global hemostasis/fibrinolysis, as evidenced by changes in functional coagulation tests –ROTEM or OHP. Secondary outcomes included the assessment of effects on conventional coagulation laboratory tests, such as D-dimer, fibrinogen, APTT, PK/INR, platelet count, antithrombin, Protein C, and Protein S. To evaluate the impact of LMWH on test results, Anti-FXa levels were compared across groups.

### Assays

All patients underwent testing for either total IgG SARS-CoV-2 antibodies or real-time reverse-transcriptase polymerase-chain-reaction (rRT-PCR) to confirm SARS-CoV-2 infection. Venous blood samples for conventional coagulations tests (CCTs) and ROTEM were collected from each participant at baseline, on day 5, and on day 29 of hospitalization. Additionally, CCTs were obtained at day 9 and day 14. An extra test citrate 3,2% tube of venous blood was drawn at each sampling occasion for plasma storage. The plasma was centrifuged at 3000 G for 10 minutes, and stored at -80°C within 3 hours after collection. These stored samples were later utilized for OHP, scanning electron microscopy (SEM), and Anti-FXa assessments.

A timeline detailing treatment administrations and blood sample testing is presented in Figure 2.

**Figure 2.**
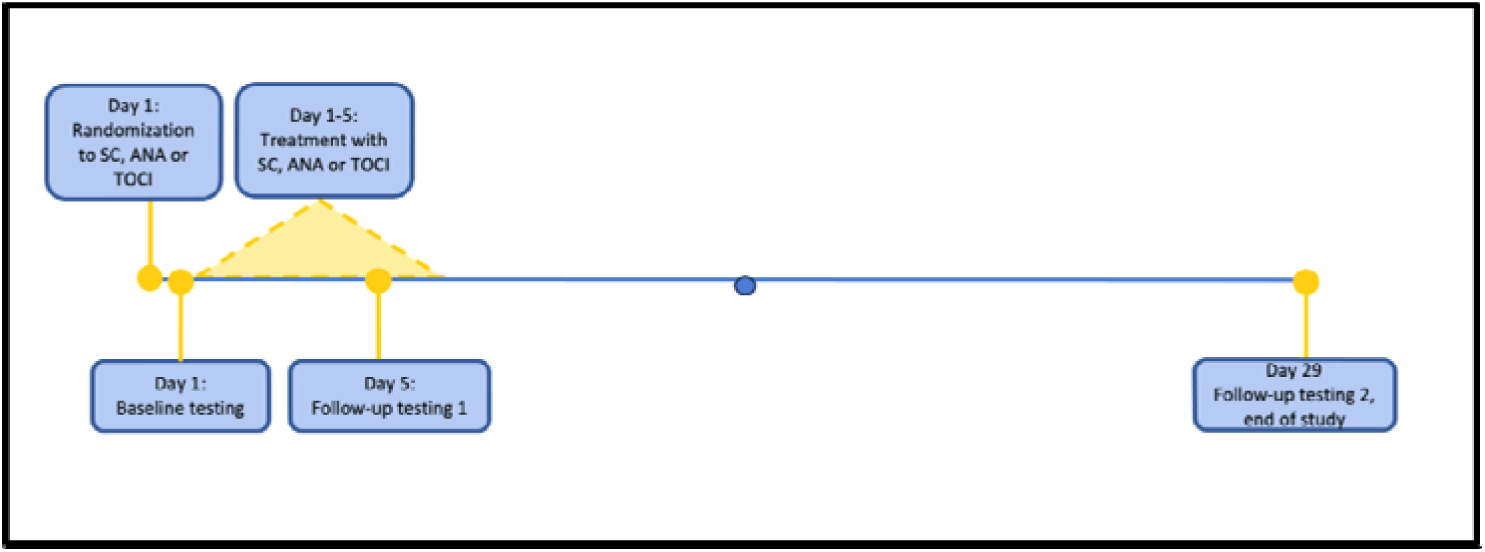
Timeline.

### Rotational Thromboelastometry (ROTEM)

ROTEM is an established whole-blood analysis method, providing insights into whole-clot formation and dissolution ^30^. This technique is instrumental in detecting and monitoring coagulopathy, offering a rapid assessment at the clinical point of care. Thromboelastometric analyses were conducted using a ROTEM sigma instrument (Tem Innovations GmbH, Germany). Three ROTEM-variables were analyzed: (1) extrinsically activated assays with tissue factor (EXTEM), (2) intrinsically activated assays using phospholipid and ellagic acid (INTEM), and (3) fibrin-based extrinsically activated assays with tissue factor and platelet inhibitor cytochalasin D (FIBTEM). EXTEM and INTEM assess the extrinsic and intrinsic pathways, respectively, while FIBTEM provides insights into fibrinogen function by eliminating platelet contribution to clot formation.

Within every ROTEM-variable, we quantified the following parameters: Clotting time (CT), which is the time (in seconds) from test start until an amplitude of 2 mm is reached, providing information about coagulation activation/initiation; Clot formation time (CFT), corresponding to the time (in seconds) between 2 and 20 mm amplitudes, offering insights into clot propagation; Maximum clot firmness (MCF), which is the maximum amplitude (in mm) reached during the test, conveying information about clot stability; and Lysis index (LI) 45, representing the percentage of remaining clot stability in relation to MCF 45 minutes after CT, expressed in percent ^31^. The coefficient of variation for CT was < 20%, and for MCF <10%.

### Overall Hemostatic Potential (OHP)

OHP serves as a global hemostatic assay employed to evaluate alterations in overall coagulation and fibrinolysis. This assay discerns changes in the clot formation process, where generated thrombin converts fibrinogen in the plasma sample to fibrin. Simultaneously, it detects variations in fibrinolysis, where plasminogen activation produces plasmin, leading to the breakdown of the formed fibrin. The methodology involves repeated spectrophotometric registrations of the fibrin-aggregation curve, with each absorbance value representing the level of fibrin in a sample at a specific time point, where the area under the curve (AUC) reflects the balance between fibrin generation and degradation over time.

In this assay, citrated plasma is recalcified and supplemented with thrombin (Sigma-Aldrich) at 0.04 U/mL, with or without tissue plasminogen activator (t-PA) (Boehringer-Ingelheim) at 300 ng/mL, resulting in the formation of two fibrin- aggregation curves. The real-time absorbance change in the sample is monitored at λC=C405Cnm every 12Cseconds for one hour. The values for the overall coagulation potential (OCP) and the overall hemostatic potential (OHP) are calculated based on the fibrin formation and fibrinolysis curves, respectively. The difference these curves provides the overall fibrinolytic potential (OFP), expressed as a percentage ^32,33^.

### Scanning electron microscopy (SEM)

SEM enables the topographical visualization and examination of biological specimens, necessitating sophisticated preparative and imaging procedures ^34^. This technique proves highly sensitive to cellular changes and fibrin structures, rendering it well-suited for visualizing clot ultrastructure ^35^. In our study, we conducted analyses on four patients from each study arm at three different time points (baseline/ day 5 /day 29). Briefly, after performing the OHP assay, the clots were washed, fixed in 2.5% glutaraldehyde, and stored at 4°C. The specimens were then analysed using an Ultra 55 field emission scanning electron microscope (Carl Zeiss), and individual fibre thickness was measured. SEM samples were blinded to the laboratory personnel. An illustrative image from each group, visualized through SEM, is presented in the article (Fig. 5).

### Data analysis/Statistics

Baseline characteristics were presented as medians and interquartile ranges (IQR) for continuous variables, and as counts and percentages for categorical variables. Summaries were provided for the entire study population as well as separately for each treatment group. Similarly, baseline values for outcome variables were reported using the same methods.

A linear mixed effects model with random intercepts was used for the main analysis. Treatment was considered a categorical exposure, interacting with time as a continuous variable. The SC treatment group served as the reference category in the model. The data indicated that some outcomes may move non-linearly over time, but due to the size of the data set larger models were unable to be fitted.

Baseline characteristics were not adjusted for, as they were not expected to influence time-dependent differences in outcome variables. Survival calculations were not feasible due to insufficient statistical power. The results from the models are presented as graphs showing how the outcomes differ over time across the three treatment groups. Only the fixed effects are presented in these graphs. The intercepts in the graphs represent estimated values within the model, and should not be confused with median values.

For the confirmatory analysis, we examined whether differences in global hemostasis assay results depended on treatment group, specifically evaluating how these differences changed over time. In the exploratory analyses, results were evaluated from conventional coagulation laboratory tests, and visualized fibrin networks using SEM. This was done in four patients from each treatment arm in order to assess and compare fibrin network density and fiber diameter between groups. Additionally, we assessed heparin effects on test results by measuring Anti-FXa activity in plasma.

This study was conducted and reported in accordance with the Consolidated Standards of Reporting Trials (CONSORT) guidelines. A preregistration was completed on aspredicted.org in April 2023. R.Studio version 4.3.1 was utilized for modeling and visualizations, and p-values below 0.05 were considered statistically significant. The code is available upon request.

## Results

Of the 74 enrolled patients, 78% were men. The median age at recruitment was 62 years, with a median BMI of 28 kg/m^2^. Additionally, 38% of the participants had diagnosed hypertension, and 20% had diabetes mellitus. The baseline characteristics and conventional coagulation test results are presented in Table 1.

**Table 1.** Baseline characteristics. N(%) / median (IQR).

| TABLE 1. | ALL<br>n=74 | SC<br>n=26 | ANA<br>n=27 | TOCI<br>n=21 | Reference interval |
| --- | --- | --- | --- | --- | --- |
| Age | 62 (54-70) | 61.5 (54-68) | 61 (53.5-71) | 64 (57-70) |  |
| Sex male | 58 (78%) | 19 (73%) | 21 (78%) | 18 (86%) |  |
| BMI | 28 (26-32) | 28 (26-30) | 28 (24-34) | 28 (27.5-32) |  |
| P-Fibrin D-dimer (mg/L) | 1.0 (0.6-1.5) | 1.0 (0.6-1.8) | 0.9 (0.6-1.3) | 1.2 (1.0-1.4) | < 0.98 |
| P-Fibrinogen (g/L) | 6.6 (5.8-8.0) | 6.1 (5.4-7.6) | 6.6 (5.8-8.2) | 7.3 (6.3-8.2) | 2.0-4.2 |
| P-APTT (s) | 24 (21-27) | 26 (22-27) | 24 (22-27) | 23 (21-27) | 20-30 |
| P-PK (INR) | 1.1 (1-1.1) | 1 (1-1.1) | 1.1 (1-1.2) | 1 (1-1.2) | ≤ 1.2 |
| Platelet count (10<sup>9</sup>/L) | 241 (200-285) | 245 (208-371) | 239 (197-273) | 247 (195-300) | Men 145-348, women 165-387 |
| P-Antithrombin (kIE/L) | 1.0 (0.9-1.1) | 1.0 (0.9-1.1) | 1.0 (0.9-1.1) | 1.0 (0.9-1.1) | 0.8-1.2 |
| P-Protein C (kIE/L) | 1.1 (0.9-1.2) | 1.1 (0.9-1.2) | 1.0 (0.9-1.1) | 1.1 (0.9-1.4) | 0.7-1.4 |
| P-Protein S (kIE/L) | 0.8 (0.6-0.9) | 0.8 (0.6-0.9) | 0.7 (0.6-0.8) | 0.9 (0.8-0.9) | Men 0.73-1.31, women 0.65-1.15 |
| B-Hemoglobin (g/L) | 131 (124-141) | 134 (122-141) | 131 (127-144) | 130 (125-136) | Men 134-170, women 117-153 |
| P-Anti-FXa (kIE/L) | 0.17 (0.06-0.45) | 0.14 (0.04-0.42) | 0.15 (0.06-0.31) | 0.31 (0.09-0.58) | < 0.10 |
| Ischemic heart disease | 5 (7%) | 1 (4%) | 2 (7%) | 2 (10%) |  |
| Chronic heart failure | 1 (1%) | 0 (0%) | 0 (0%) | 1 (5%) |  |
| Peripheral arterial disease | 2 (3%) | 0 (0%) | 0 (0%) | 2 (10%) |  |
| Previous CVL | 5 (7%) | 3 (12%) | 2 (7%) | 0 (0%) |  |
| Hypertension | 28 (38%) | 10 (38%) | 11 (41%) | 7 (33%) |  |
| Diabetes | 15 (20%) | 5 (19%) | 5 (19%) | 5 (24%) |  |
| Asthma/COPD | 8 (11%) | 3 (12%) | 3 (11%) | 2 (10%) |  |

*Table 2* displays the results of global hemostasis tests at baseline. Even though this study was originally designed as a randomized trial, certain laboratory test results exhibited variations among treatment groups at baseline (see Limitations section).

**Table 2.**
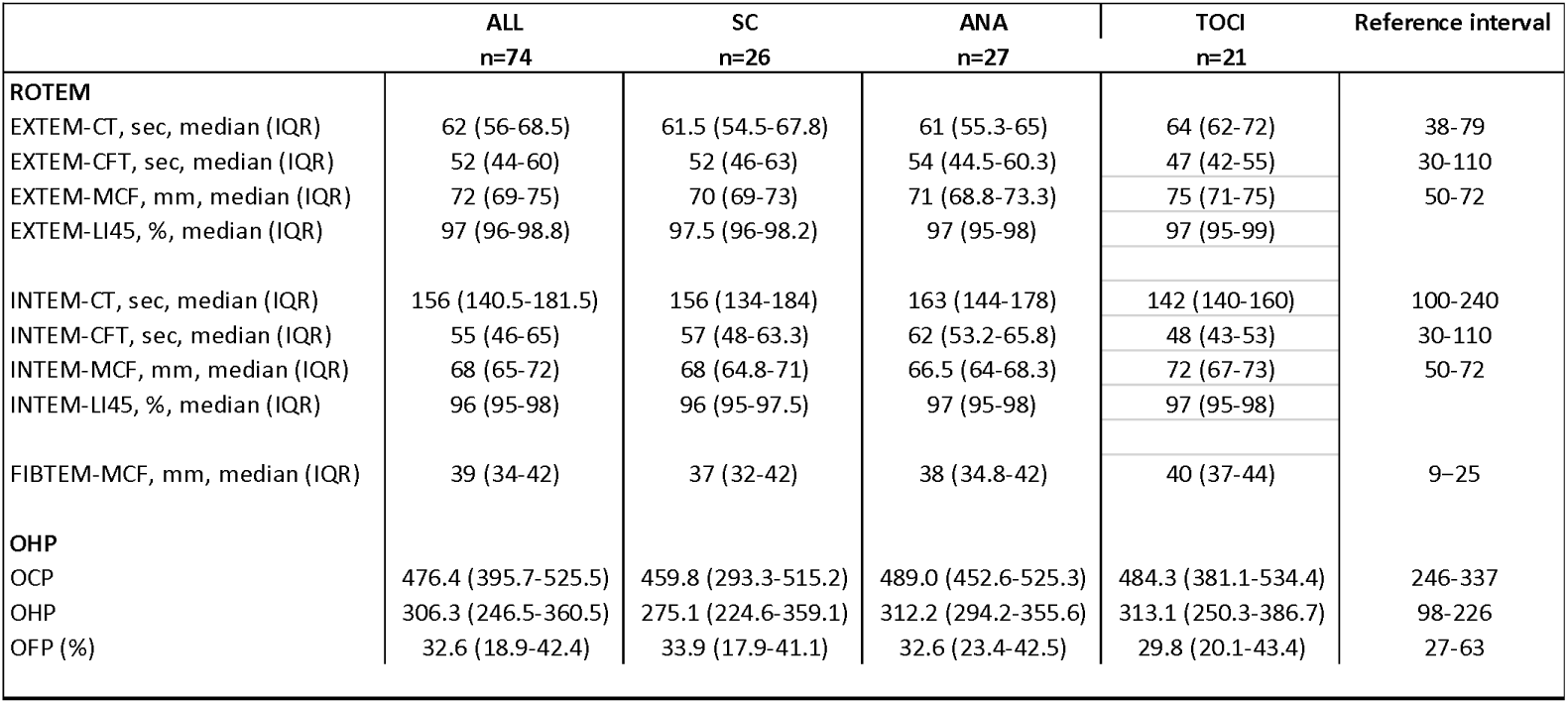
Baseline levels of ROTEM and OHP data. Median (IQR).

### Primary outcomes

#### ROTEM

At baseline, FIBTEM-MCF exceeded the reference range in all treatment groups, indicating hypercoagulation. However, all other ROTEM variables were within the reference range at the study’s initiation.

Over time, ROTEM variables reflecting coagulation activation, specifically EXTEM-CT and INTEM-CT, demonstrated a prolongation, indicating decreasing hypercoagulation tendency. The increase in EXTEM-CT was notably more pronounced in the ANA group compared to SC (*p*=0.036). No significant differences were observed between TOCI and SC, or between TOCI and ANA treatments (Fig. 3a). Regarding INTEM-CT, increasing levels were associated with LMWH treatment and Anti-FXa levels (Fig. 6d).

**Figure 3.**
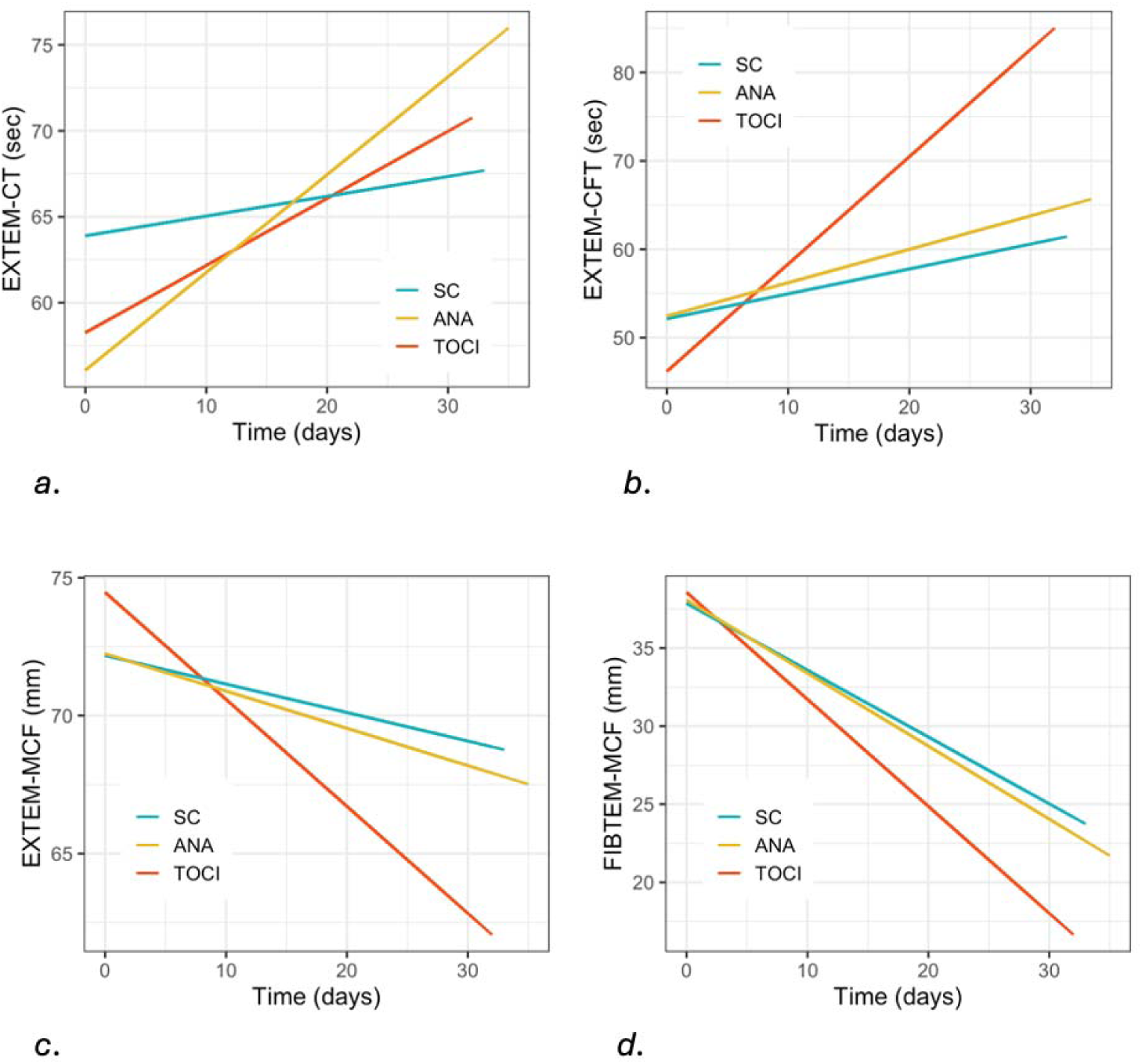
Linear mixed model graphs of ROTEM variables. a) EXTEM-CT, b) EXTEM-CFT, c) EXTEM-MCF and d) FIBTEM-MCF.

EXTEM-/INTEM-CFT, representing the time for clot propagation, increased over time. The increase was significantly more pronounced in the group of patients receiving tocilizumab (EXTEM-CFT *p*<0.001) both compared with patients receiving SC, and compared with patients receiving anakinra (Fig. 3b). No discernable difference was observed between ANA and SC groups.

EXTEM-/INTEM-/FIBTEM-MCF, indicating clot firmness, were reduced over time. The reduction was significantly more pronounced in the TOCI group (EXTEM-MCF *p*<0.001, FIBTEM-MCF *p*=0.005) compared with ANA as well as compared with SC (Fig. 3c-d). No difference was seen between ANA and SC groups.

ROTEM Lysis Tests: In the ROTEM lysis tests (EXTEM-/INTEM-LI30, EXTEM-/INTEM-LI45) measuring fibrinolysis in %, high levels were observed. No significant differences were observed between treatment groups over time.

#### OHP

OHP variables exhibited increased OCP at baseline compared with healthy controls ^33^ (*p*=0.08), diminishing over time in all treatment groups, with no significant differences observed between treatment groups. The OHP levels decreased and OFP levels increased over time in all treatment groups, with no significant differences between groups (Fig. 4 a-c). OHP and OFP did not differ significantly at baseline (*p*=0,27; *p*=1) from healthy controls ^33^.

**Figure 4.**
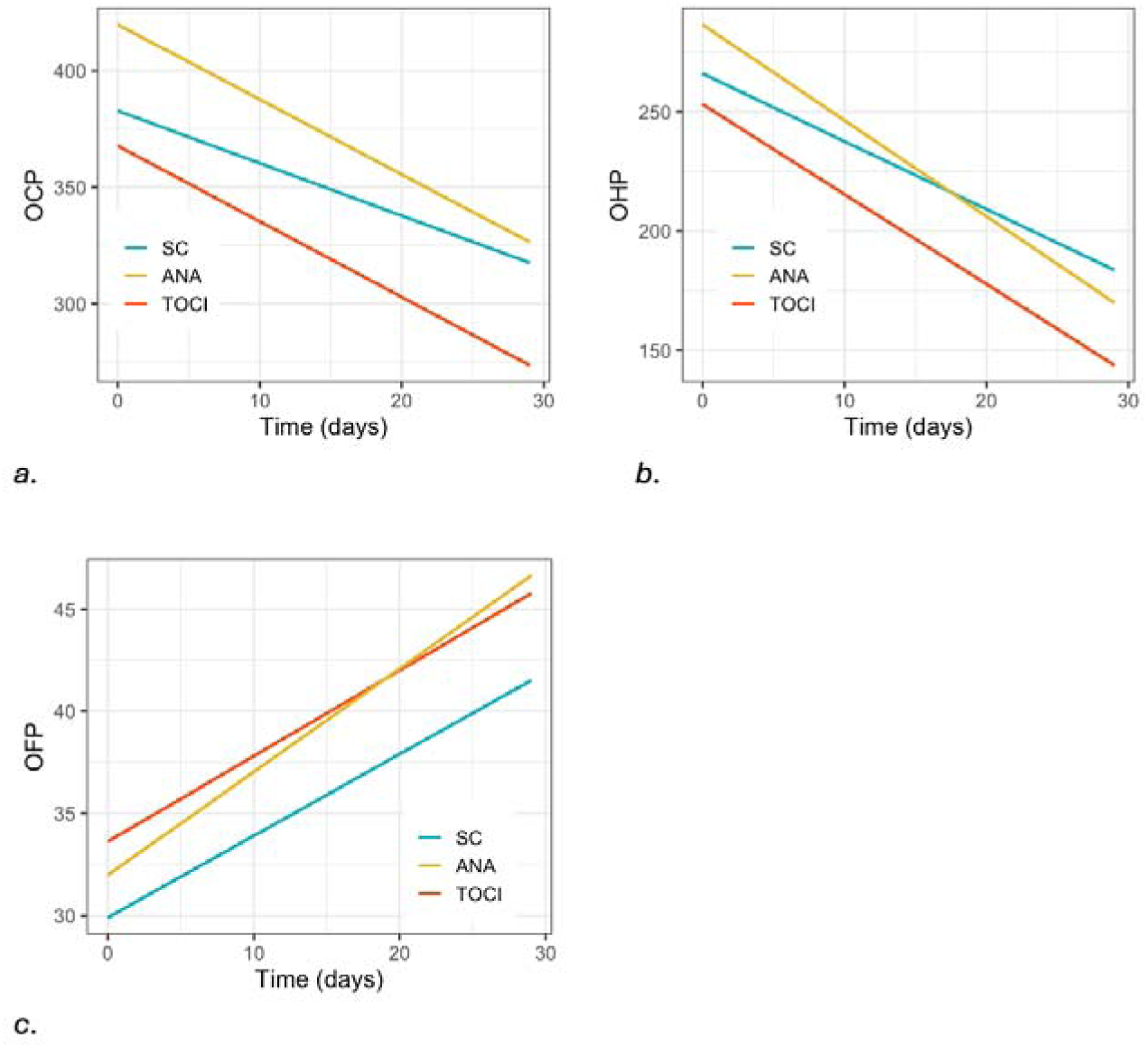
Linear mixed model graphs of OHP variables. a) OCP, b) OHP and c) OFP.

#### SEM

Three images from each treatment group, captured at baseline/day 5/day 29, visually illustrate clot structural changes over time (Fig. 5 a-i). Figure 5 a-c exemplifies patients in the SC-group at baseline (Fig. 5a), day 5 (Fig. 5b) and day 29 (Fig. 5c), figure 5 d-f demonstrates patients in the ANA group at baseline (Fig. 5d), day 5 (Fig. 5e) and day 29 (Fig. 5f), and figure 5 g-i represents patients in the TOCI group at baseline (Fig. 5g), day 5 (Fig. 5h) and day 29 (Fig. 5i).

**Figure 5.**
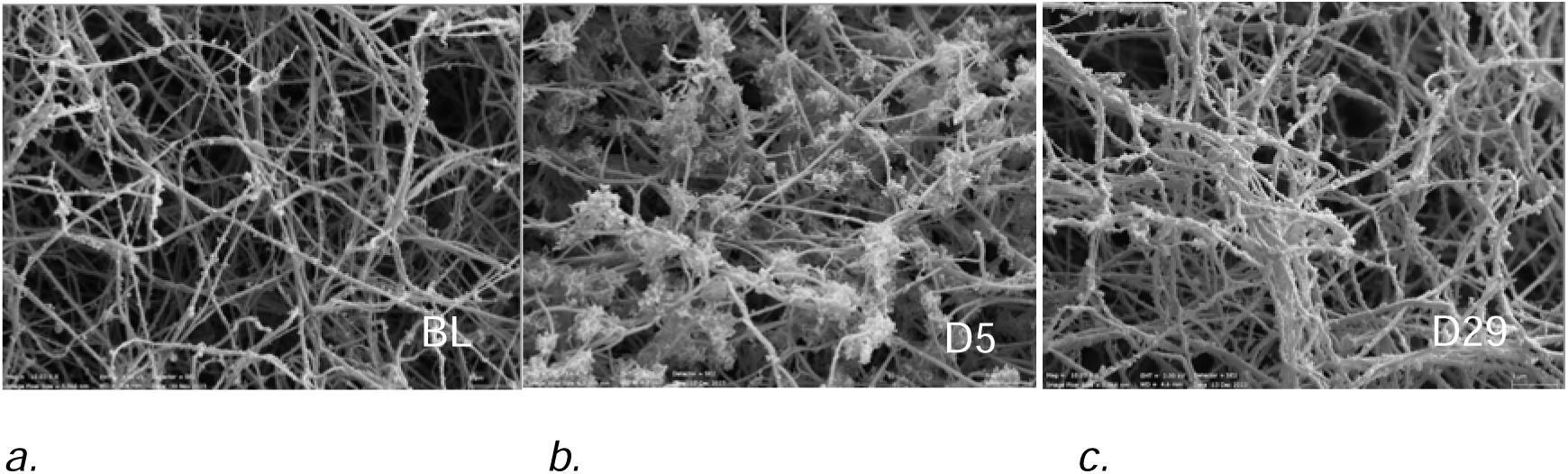
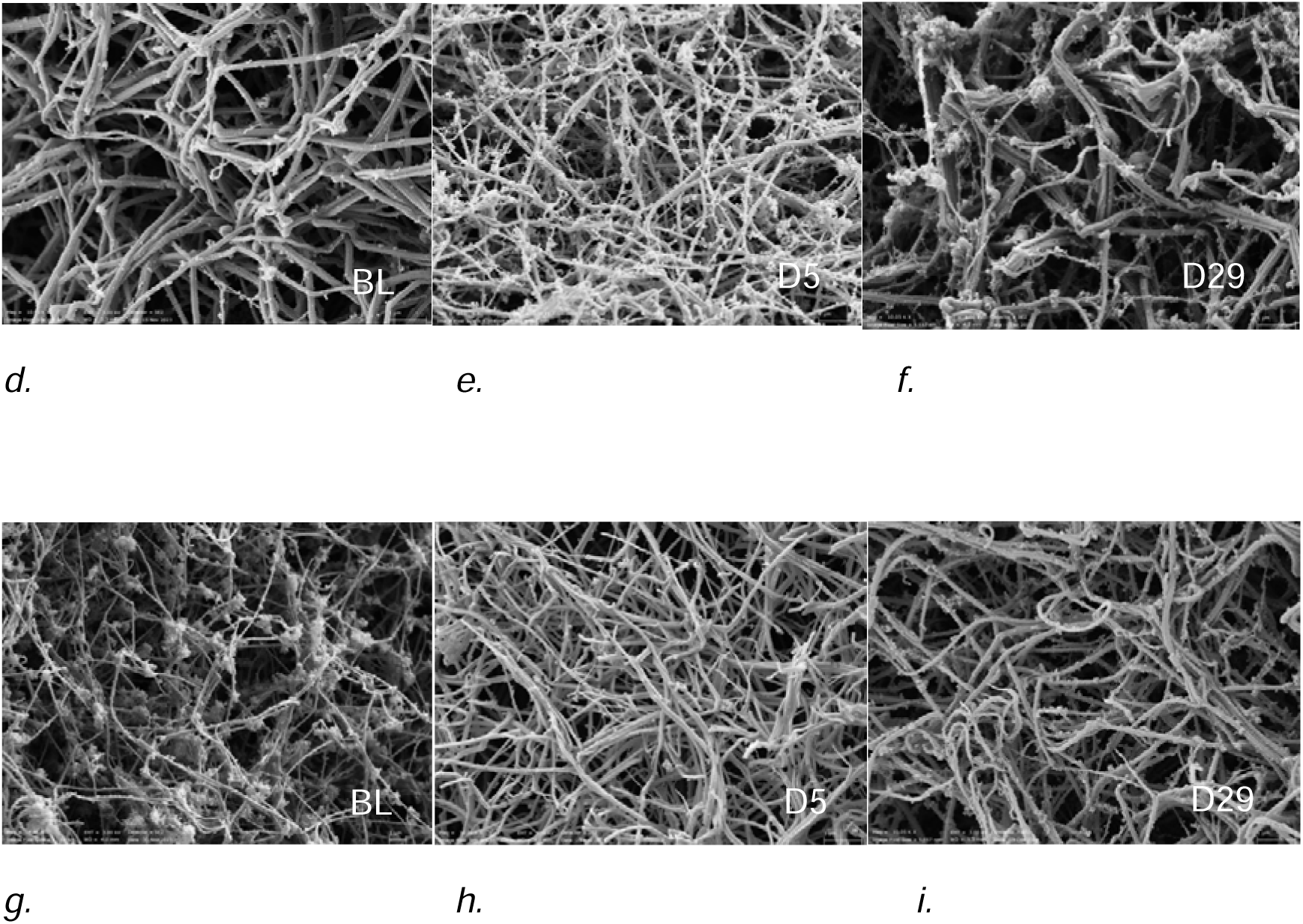
SEM in patient in SC treatment group at a) baseline b) day 5 and c) day 29. SEM in patient in ANA treatment group at d) baseline e) day 5 and f) day 29. SEM in patient in TOCI treatment group at g) baseline h) day 5 and i) day 29.

From baseline to day 5, fibrin network structures exhibited increased density in all treatment groups. In contrast, between day 5 to day 29, the ANA and TOCI groups showed indications of remodeling in the fiber clot structure. This restructuring involved a decrease in fiber density, the emergence of thick fibrin fiber branches from thinner individual fibers, and the presence of larger pores.

### Secondary outcomes

#### CCTs

D-dimer levels were elevated at baseline (*p*=0.004) and decreased over time in the study period across all treatment groups, aligning with the observed diminishing hypercoagulation tendency in global coagulation tests. While there was no significant difference between TOCI and SC over time, the drop in D-dimer levels was significantly less pronounced in the ANA group compared with SC (*p*=0.048) There was no difference observed between the ANA and TOCI groups (Fig. 6a).

**Figure 6.**
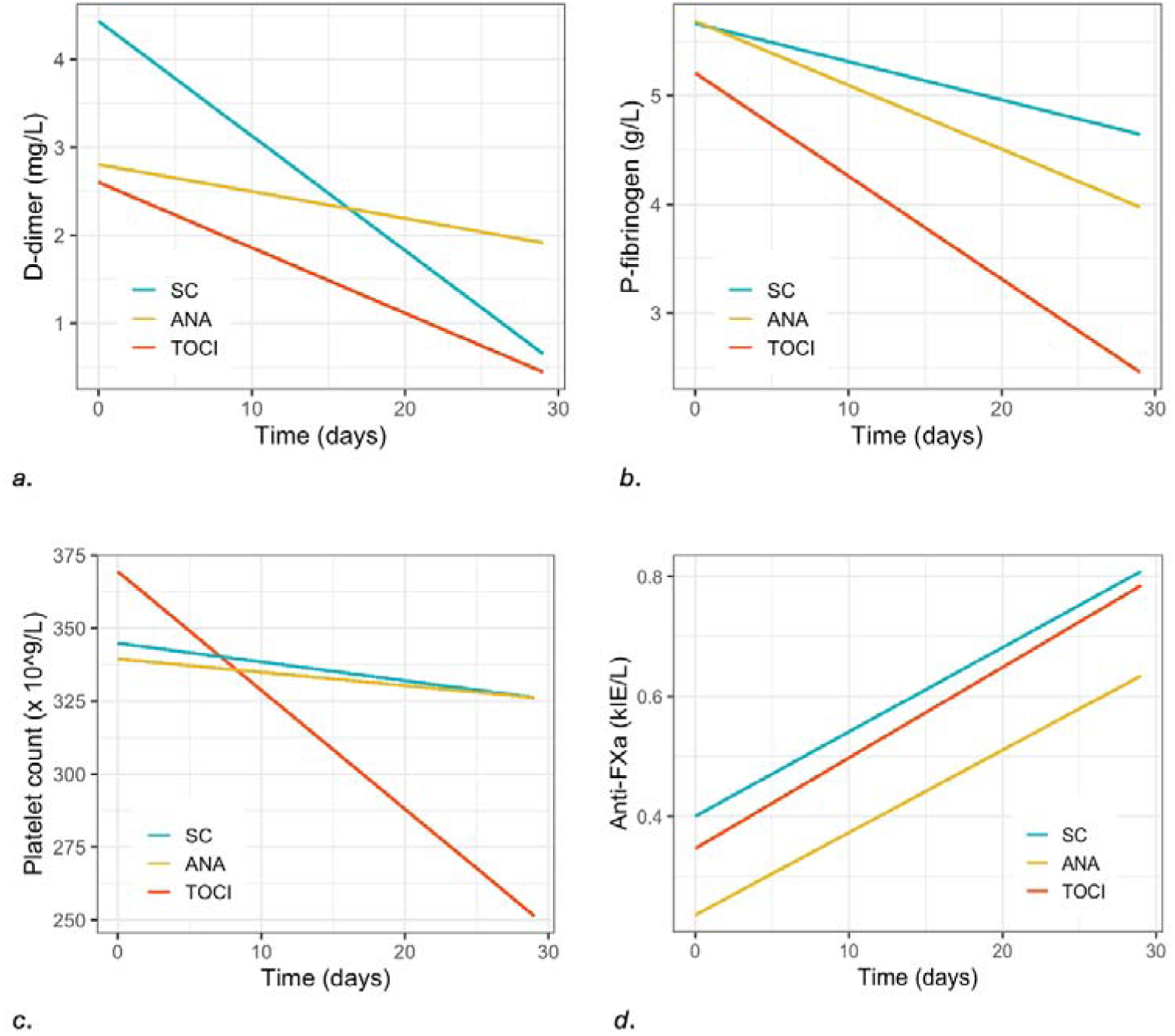
Linear mixed model graphs of CCT variables. a) D-dimer, b) P-Fibrinogen, c) Platelet count and d) Anti-FXa.

Fibrinogen levels were elevated at baseline and declined over time in all groups when tested in a linear mixed model. The decrease over time was more pronounced in the TOCI group (*p*=0.004) compared to the SC group. There were no significant differences observed in the decrease over time between the ANA group and SC group, or in the ANA group compared with TOCI (Fig. 6b).

Platelet count levels were within reference range at baseline and decreased over time in all groups, remaining within reference levels at the end of the study period. However, the decrease was more pronounced in the TOCI group (*p*=0.007) both compared with ANA and SC. There was no difference observed between ANA and SC (Fig. 6c).

To assess confounding effects of LMWH treatment on test results, trough concentrations of anti-FXa were measured. Anti-FXa levels increased over time, with no significant differences between groups. (Fig. 6d).

## Discussion

Whereas COVID-19 infection is associated with profound changes in the coagulation system, involving clot strength and fibrin networks, this clinical study provides evidence of a reduced hypercoagulation tendency in global test results among hospitalized COVID-19 patients receiving tocilizumab in addition to standard care, compared with patients receiving anakinra in addition to standard care, or standard care alone.

### ROTEM

The effects on global coagulation tests were more pronounced in the TOCI group after 29 days. This was evident through the prolongation of EXTEM-/INTEM-CT and EXTEM-/INTEM-CFT, along with a decrease in EXTEM-/INTEM-/FIBTEM-MCF levels. These results suggested clot activation/propagation rates and clot firmness closer to the lower part of the reference intervals, indicating a less hyperactive coagulation system in patients receiving tocilizumab in addition to standard care for COVID-19, as opposed to patients receiving anakinra or standard care alone.

### OCP/OHP

OHP data indicated elevated OCP levels at baseline, reflecting a heightened fibrin generation at the start of the study, which gradually decreased during the time of hospitalization. Consistent with this, Lee et al. presented evidence of significantly elevated fibrin generation (characterized by high OCP and OHP levels) and impaired fibrinolysis (indicated by reduced OFP levels) in COVID-19 patients compared with healthy controls, suggesting dysregulation between the coagulation and fibrinolytic pathways ^20^. Unlike the ROTEM results, no significant differences were observed between treatment groups in OHP test results. The dissimilarity may be attributed to variations in analysis materials; OHP utilizes platelet-poor plasma, while whole blood is used in ROTEM. As validated by Pretorius et al., who compared electron microscopy images of hypercoagulability in platelet-poor plasma versus whole blood, more rigid clot structures were evident in whole blood images, where pseudopodia were extended and merged into the fibrin network^35^.

### Fibrinolysis

The fibrinolytic activity indicator OFP was in the lower part of the reference interval, increasing over time. In the ROTEM lysis tests, high levels were observed, indicating low fibrinolytic activity. However, no significant differences were observed between groups in ROTEM lysis test results over time. This aligns with earlier studies indicating reduced fibrinolysis in COVID-19 patients, suggesting an imbalance in procoagulative and fibrinolytic mechanisms contributing to the disease’s complex pathophysiology ^20,36–38^.

### SEM

SEM images revealed alterations in fibrin network structures and clot morphology over time, with discernible differences in the ANA and TOCI treatment groups compared with the SC group. This observation is further substantiated by the gradual increase in porosity and sparser distribution of fibers within the fibrin network structures of the ANA and TOCI treatment groups from day 5 to day 29, as opposed to the SC group. These findings suggest a reduction in hypercoagulation tendencies within these groups during the mentioned timeframe, contributing to a differentiated branching process and a less densely packed matrix. This trend was visually less pronounced in the SC group. Consequently, SEM images align with and support our findings of a diminishing hypercoagulation tendency in the TOCI treatment group over time. The resemblance of clot structures between ANA and TOCI groups as observed in SEM images, contrasts with their distinct outcomes in the statistical analyses of our study, where ANA in most analyses did not differ from SC. However, it is important to recognize, that laboratory tests and clot structure images serve different purposes, capturing different aspects of a condition, where visual interpretations from images may not always correlate directly with findings from blood samples.

It is widely recognized that an increased tendency for fibrin production is a fundamental characteristic in moderate and severe COVID-19. This arises due to heightened procoagulant activity, coupled with elevated fibrinogen levels and diminished fibrinolysis ^39^. Our findings indicate a diminished (though non-significant) OFP in the SC group, along with higher fibrinogen levels at all time points compared with the ANA and TOCI groups. This may partly account for the observed trend of fibrin network structures becoming progressively more porous, and fibers more sparsely distributed, in the ANA and TOCI treatment groups from day 5 to day 29, as opposed to the SC group. However, no significant difference was noted between the ANA and TOCI groups. It is conceivable that in COVID-19, the polymerization process of the fibers may not directly correspond to the treatment with interleukin inhibitors, thereby emerging as a late-stage structural determinant. Nonetheless, further mechanistic studies are warranted to delve into this aspect.

### P-fibrinogen

Within our dataset, baseline P-fibrinogen levels indicated hypercoagulation. Notably, patients subjected to IL-6 treatment exhibited a significant reduction in P-fibrinogen levels over time compared with other treatment groups, affirming a diminished hypercoagulative state with decreased clot density in the IL-6 group. This aligns with prior data suggesting that IL-6 plays a regulatory role in P-fibrinogen levels in the blood, stimulating hepatocytes to produce fibrinogen in a dose-dependent manner ^40^. Furthermore, the inhibitory effect of tocilizumab on the binding of IL-6 to its receptor is associated with a significantly reduction in P-fibrinogen levels ^41^.

### Platelet count

Thrombocytopenia (defined by a platelet count < 150 x 10^9^/L) upon hospital admission due to COVID-19, is strongly associated with poor outcomes and mortality, and is attributed to increased platelet activation and consumption ^42^. In our dataset, 5 patients (7%) exhibited thrombocytopenia at baseline, and of these, 4 had levels > 150 x 10^9^/L by day 29. Mean platelet count levels, although remaining within the reference range, demonstrated a significant decrease over time from baseline in the IL-6 group compared with SC, with no discernible differences compared to the IL-1 group.

Decreasing platelet counts are believed to be associated with a combination of reduced inflammatory burden and a state of platelet consumption, underscoring the crucial role of platelets in immune responses and the orchestration of inflammation. As key participants in COVID-19, platelets directly interact with the SARS-CoV-2 virus, leading to platelet hyperactivation. Due to their composition, which includes phospholipids and other components involved in the coagulation process, the surface properties of platelets are important in reinforcing clot rigidity by regulating fibrin polymerization ^43,44^.

### D-dimer

D-dimer levels exhibited an increase at baseline but decreased over time across all groups. The observation of elevated D-dimer levels, together with the presence of fibrin deposits in the lungs of COVID-19 patients, implies that fibrin-dissolving pathways are active and functional. However, there seems to be an imbalance with the fibrin-forming (thrombin-generation) system, indicating an insufficiency to counterbalance the enhanced procoagulant activity ^36,45^.

### Anti-FXa

Anti-FXa levels increased over time, with no significant differences between groups. Increasing anti-FXa levels may either indicate that higher doses of LMWH treatment were administrated at the end of study compared to baseline, or that patients experienced renal failure, leading to impaired renal clearance of LMWH. Adjustment for anti-FXa levels was not made in the model, since we did not suspect them to influence changes in test results over time.

The favorable outcome of IL-6 antagonists in our data, indicating a reduction in hypercoagulation properties over time, align with previous studies. Kovács et al noted that elevated blood levels of IL-6 led to thrombocytosis, platelet hyperactivity, and platelet aggregation ^25^. Furthermore, Hou et al demonstrated that IL-6 not only increased platelet count, but also influenced platelet function by enhancing their responsiveness to thrombin stimulation ^40^.

Together, the observed impacts of IL-6 antagonists on coagulation profiles in our data suggest a close link to the survival benefits of tocilizumab in severe COVID-19. To determine whether these effects stem from diminished coagulation/inflammation activity or direct interleukin antagonist effects, further randomized studies are warranted.

## Limitations

This study has limitations. Firstly, as the original ImmCoVA study was discontinued due to national changes in standard care, our data are underpowered for survival calculations. Secondly, despite adherence to a randomized study protocol, there were variations in baseline values among treatment groups, specifically in EXTEM-CT, with ANA differing from SC (*p*=0. 048). Thirdly, the sample size of 74 patients was relatively small, posing the risk of a type 2 error. Fourth, all data were collected at a single site, limiting its external validity. Fifth, due to the linear mixed model design, an accurate visualization of non-linear outcome changes is challenging.

Sixth, the SEM results are constrained by their reliance on visual confirmation, introducing a limitation to the evaluation process in assessing fiber structures. Lastly, not all patients were included in the SEM analysis.

## Conclusion

In patients with COVID-19 receiving the interleukin-6 antagonist tocilizumab in addition to standard care, global hemostasis tests and conventional coagulation laboratory tests were showing less signs of hypercoagulation after 29 days, compared with patients receiving the interleukin-1 antagonist anakinra or standard care alone. Tocilizumab significantly diminished hypercoagulation by increasing clotting time, clot formation time, and reducing clot strength, confirming direct effects of IL-6 antagonists on the coagulation system in severe COVID-19.

## Data Availability

All data produced in the present study are available upon reasonable request to the authors

## Acknowledgements

We extend our gratitude to the staff at the Laboratory Unit of Karolinska University Hospital, particularly Nida Mahmoud, for their invaluable assistance in conducting the blood samples. This study was funded by a grant from the Swedish Research Council (2020-06318 JL;JSC). The funder had no role in study design, data collection and analysis, decision to publish, or preparation of the manuscript.

